# Community-level variation in TB testing history: analysis of a prevalence survey in Blantyre, Malawi

**DOI:** 10.1101/2023.04.28.23289249

**Authors:** Emily S Nightingale, Helena R A Feasey, McEwen Khundi, Rebecca Nzawa Soko, Rachael M Burke, Marriott Nliwasa, Hussein Twabi, James A Mpunga, Katherine Fielding, Peter MacPherson, Elizabeth L Corbett

**Affiliations:** London School of Hygiene and Tropical Medicine, London, UK; Malawi Liverpool Wellcome Trust Clinical Research Programme, Blantyre, Malawi; University of Malawi College of Medicine, Zomba, Malawi; National TB Programme, Government of Malawi, Malawi; Liverpool School of Tropical Medicine, Liverpool, UK; University of Glasgow, Glasgow, UK

## Abstract

Equitable access to tuberculosis testing is vital for achieving global treatment targets, but access to diagnostic services is often worse in poorer communities. The SCALE survey to estimate TB prevalence in Blantyre city, Malawi, also recorded engagement with TB services. We explored variation in self-reported TB testing history between 72 community clusters - adjusting for sex, age and HIV status - and investigated whether residual differences could be explained by household poverty. We observed substantial variation between clusters in the prevalence of ever-testing for TB, with little correlation between neighbouring clusters. Participants in poorer households had, on average, lower odds of ever-testing, yet adjusting for poverty did not reduce cluster-level variation. We conclude that, despite a decade of increased active case finding efforts, access to TB testing is inconsistent across the population of Blantyre, likely reflecting health inequities that also apply to TB testing in many other settings.

## Background

Tuberculosis (TB) was second only to COVID-19 as an infectious cause of death in 2021, with 1.6 million deaths globally. Every year many people develop active TB disease but either do not receive a diagnosis, or are not registered as having been linked to care. In 2021, an estimated 42% of people who developed TB disease did not receive care, equivalent to 1.5 million people (1).

Suboptimal performance and high cost of available diagnostic tests present a major challenge to timely diagnosis and patient care, with available tests being difficult to decentralise and often require multiple health clinic visits. This in turn leads to high direct and indirect costs of case-seeking (2,3) before receiving a diagnosis, compounded by difficulties in accessing primary care. Poor or otherwise vulnerable populations may then have particular difficulties in accessing TB testing, motivating a re-focus of interventions to target these particular groups (4).

Most countries rely on TB case notification rates (CNRs), based on annual TB registrations and population estimates, but estimates based on CNRs alone are vulnerable to underascertainment, whereby falling TB CNRs may represent either public health success - with reduced transmission and new disease incidence - or failure to diagnose and notify. TB prevalence surveys (5) for undiagnosed active TB disease provide an independent assessment of TB burden, allowing calculation of prevalence-to-notification ratios that measure how well the health system is performing with respect to TB testing and diagnosis (6).

National TB prevalence surveys are typically powered to provide precision in TB estimates within one to two percentage points at a national level. Data on *sub*national TB prevalence - needed to produce meaningful targeted public health action - are often scarce. Spatially targeted screening guided by such data has been effective in reducing transmission in some low-burden settings, but remains to be fully explored in high-burden settings such as Malawi (7).

In Malawi, we conducted the Sustainable Community-wide Active case-finding for Lung hEalth (SCALE) study in 2019-2020 (8). This combined a city-wide TB prevalence survey using digital chest radiography, symptom screening and bacteriological tests on sputum if indicated in 72 high density residential neighbourhoods with a questionnaire, including questions about previous TB testing behaviour. Although power was limited by lower than anticipated prevalence of bacteriologically confirmed TB (28 previously undiagnosed patients, and 1 already on treatment), we combined these with case-notification data to identify neighbourhoods with high prevalence-to-notification ratios consistent with substantially delayed/underdiagnosed TB (6).

Unlike for HIV, data on past TB testing are not routinely reported to WHO, or included in demographic health surveys. (5). As such, relatively little is known about how testing varies by demographic characteristics or geographical location. We investigated neighbourhood- and household-level predictors of self-reported past TB testing, aiming to estimate how much variation in TB testing was not explained by variation in individual characteristics such as age, sex and HIV status and might, therefore, reflect structural factors (as discussed in (4)). This follows previous studies in Malawi suggesting that underdiagnosis is highest in peripheral, informal neighbourhoods that tend to be the least affluent (5,6,10,11).

TB is a disease of poverty, and providing equitable access to testing services is vital for reaching End TB goals (12,13). The main aims of this study were to quantify differences in past TB testing across Blantyre city, and to provide an initial assessment of whether or not this pattern was similar to our previous estimates of underdiagnosis based on prevalence/CNR ratios. If so, then the prevalence of self-reported past TB testing could serve as a useful alternative to costly TB prevalence surveys for identifying underserved populations likely to benefit from targeted interventions such as active case-finding interventions.

## Methods

### Setting

In Malawi, testing for TB is primarily offered through primary healthcare centres. Individuals presenting at clinics with symptoms consistent with TB are asked for a sputum sample to be tested with Xpert (14). In addition to screening within primary care, the WHO recommends routine screening for TB among people living with HIV (PLHIV; primarily delivered through ART services) and close contacts of TB cases (11). Primary care centres are not equipped to diagnose TB through chest x-ray; only the central Queen Elizabeth hospital has such facilities. However, mobile x-ray vans are used to bring this type of screening to less accessible communities, with an aim to reach individuals who may not otherwise seek care.

### Study Design

The SCALE TB prevalence survey, conducted between May 2019 and March 2020, aimed to estimate the prevalence of pulmonary TB disease among adults aged 18 years or older living in 72 community clusters in urban Blantyre, Malawi (See (8) for a more detailed description). The population within these clusters was enumerated through a census conducted in 2015 by the Malawi-Liverpool-Wellcome (MLW) research centre, and extrapolated to the period of interest using national estimates of population growth (previously described in (6)).

Households within clusters were selected at random and approached by trained survey staff to record the age and sex of all household residents, as well as a range of socio-economic indicators at the household level. All adult (≥18 years) residents were then asked to complete an individual questionnaire and were invited to attend a nearby temporary community facility for TB and HIV screening, located in the centre of the neighbourhood cluster. *Supplementary Materials A* contains a discussion of the representativeness of the sampled population and a comparison of age-sex distributions is shown in *Supplementary figure S1*.

A randomly-selected subset of 20% of participating adults were invited to complete an extended survey, which included questions about their previous experience with TB testing. Participants were asked whether they had ever submitted a sputum sample through the routine health service or community-based screening programmes, or had undergone a chest X-ray.

Participants reporting having ever undergone either form of TB testing were further asked the month and year of their last test.

### Primary outcome

The primary outcome was self-reported past TB testing, defined as having ever given a sputum sample or having ever undergone a chest X-ray (assumed to be primarily for TB diagnosis in this population).

Where the date of the last test was reported, time since the last test and testing within the last twelve months were summarised as secondary outcomes. All days were imputed as the first of the month. If only the year was reported, the month was imputed as June. If neither month nor year was reported, the date was defined as missing.

### Covariates

#### Baseline adjustment

Variation in age, sex and HIV status was accounted for by their inclusion as baseline covariates in all analyses, to explore residual trends across clusters *not* attributable to these factors. This was important because the probability of ever testing is dependent on age, and testing may further be influenced by WHO policies on screening for TB among PLHIV. By inspection of exploratory plots, age was grouped into < 25, 25-34, 35-44, 45-54 and ≥ 55 years. See *Supplementary Materials B* for a description of how participant HIV status was defined from self-reported and measured outcomes during the survey.

#### Proxy Means Test (PMT) score of poverty

The household questionnaire included a proxy means test for poverty, based on assets, food security, head of household education and sleeping conditions. (See *Supplementary Materials C* for full definitions of all constituent variables and their weighting). Previous work optimised these questions for urban Blantyre to create a composite measure of poverty predictive of the probability of living below the poverty line of $2 per day, as estimated by the Malawi 2016/17 National Integrated Household Survey (15). A logit-scale score was obtained for each household, with higher values reflecting a higher probability of poverty. For our purposes, the score was negated (such that higher score implies higher wealth) and categorised into six quantiles to align with the self-assessed measure of wealth described below.

#### Self-assessed wealth

Respondents were also asked to position themselves on a six-step scale of wealth from poorest (Level 1) to richest (Level 6), using a question (*“Imagine six steps, where on the bottom, the first step, stand the poorest people, and on the highest step, the sixth, stand the rich. SHOW THE PICTURE OF THE STEPS. On which step are you today?”*) and figure (see *Supplementary figure S2*) taken from the National Integrated Household Survey), open to their own interpretation and frame of reference. Preliminary steps were taken to validate this measure against the PMT score and to explore how the two vary spatially.

### Descriptive

We summarised the distribution of age and sex within the extended survey sample, and compared this to total population estimates of the same geographic clusters (described previously (6)), to assess the extent of bias.

We summarised the percentages of respondents reporting ever testing and testing within the last 12 months overall and by the baseline and exploratory variables of interest. The distribution across clusters was also stratified by type of testing (sputum or chest X-ray).

### Main analysis

We fitted a Bayesian logistic regression model to the outcome of having ever been tested for TB, adjusting for - at the individual-level - age group, sex and HIV status. Independent and identically-distributed (IID) random intercepts were fitted across clusters to account for correlation between individuals residing in the same neighbourhood. See *Supplementary Materials C* for a detailed model specification.

Evidence of interaction between age and sex was assessed graphically and then formally by the significance of an interaction term in the baseline model, defined by exclusion of zero from 95% credible intervals of the posterior distribution of coefficients. Model fit and convergence was assessed by Rubin-Gelman 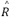 statistics, inspecting trace plots and through plots of posterior predictive checks (16,17).

The structure of variation in the fitted random effects was explored across neighbouring clusters and with respect to measures of poverty at the household level. First, evidence of spatial autocorrelation between clusters was assessed by comparing Moran’s I statistic calculated across the fitted intercepts to 999 monte-carlo simulations under the assumption of spatial independence (18). Due to the spatial isolation of some clusters, neighbours were defined by proximity of the nearest cluster centroids, as opposed to the sharing of borders. Measures of household poverty were then in turn added to the model and their relative impacts on model fit compared, with respect to leave-one-out cross-validation information criterion (LOOIC) (19) and estimated standard deviation of cluster-level random intercepts.

### Sensitivity analyses

Our primary results are from a complete case analysis. We also report models based on multiple imputation of the missing values as a robustness analysis. Five imputed datasets were created using a classification tree approach via the R package *mice* (20). Included variables were age, sex, HIV, poverty score, self-assessed wealth, knowing someone who has been tested or begun treatment for TB in the last 12 months, knowing someone who has died from TB, and ever having been tested.

## Availability of data and materials

All data and code used in this analysis are accessible via the following repository: https://github.com/esnightingale/tb-testing-history.

## Results

### Descriptive

#### Survey sample

The estimated adult population of the 72 survey clusters was 613,000. The total number of residents reported across 7175 randomly sampled households was 20,555, of whom 15,897 (76%) responded to the primary survey. 2738 (17%) respondents additionally responded to the extended questionnaire, representing 2278 unique households (*Supplementary Figure S3*). The vast majority of extended survey respondents were the only one to participate within their household (1885 households with one observation; 59 households with three or more observations). As such, adjustment for potential clustering in the outcome by household was not considered necessary.

#### Missing data

Age was unknown for three respondents (0.01%). These were assumed to be missing completely at random and excluded from complete case analysis. HIV status was classified as unknown for 117 respondents (4.3%). The rate of missingness for HIV status was higher among men than women (6.2% vs 3.0%) and among the youngest and oldest age groups (5.7% and 7.6% vs 2.2% among those aged 25-34, respectively). A further 29 respondents were missing a corresponding household survey, including the two measures of poverty. Overall, 148 (5.4%) of observations were missing some relevant covariate information.

#### Testing history by individual

Of 2,590 respondents with non-missing data, 414 (15.9%) reported having undergone some prior investigation for TB: 257 (9.9%) had given a sputum sample for testing; 262 (10.1%) had undergone a chest X-ray and 105 (4.1%) had done both. Responses for previous testing by sputum, X-ray or either were completely recorded for all respondents; however, 248 (60.0%) of those who responded yes to ever testing did not provide any information on the date of their last test. Where the date was known, this varied from less than a month to over twenty years prior to the interview, with a median of 7.2 months. Stratifying by type of TB testing, the median time since the last test was shorter for chest X-ray compared to sputum testing (6.4 vs 10.1 months, respectively). A total of 106 (4.1%) respondents reported having undergone TB testing within the last twelve months.

Reported TB testing history was similar amongst men and women and was - as would be expected - more common among older people (*Table 1*). A substantial difference was observed in testing between HIV-negative and HIV-positive individuals undergoing antiretroviral therapy (ART) (295/2,292, 12.9% vs 116/292, 39.7%), with HIV positive individuals *not* on ART falling between (8/37, 21.6%). The latter group may include newly-diagnosed or otherwise unregistered people who may have not (yet) been reached by routine TB screening and hence may have a different probability of prior testing. However, due to the small sample size of this group, all HIV positive individuals were combined for analysis, regardless of whether they were known to be on ART.

**Table 1:**
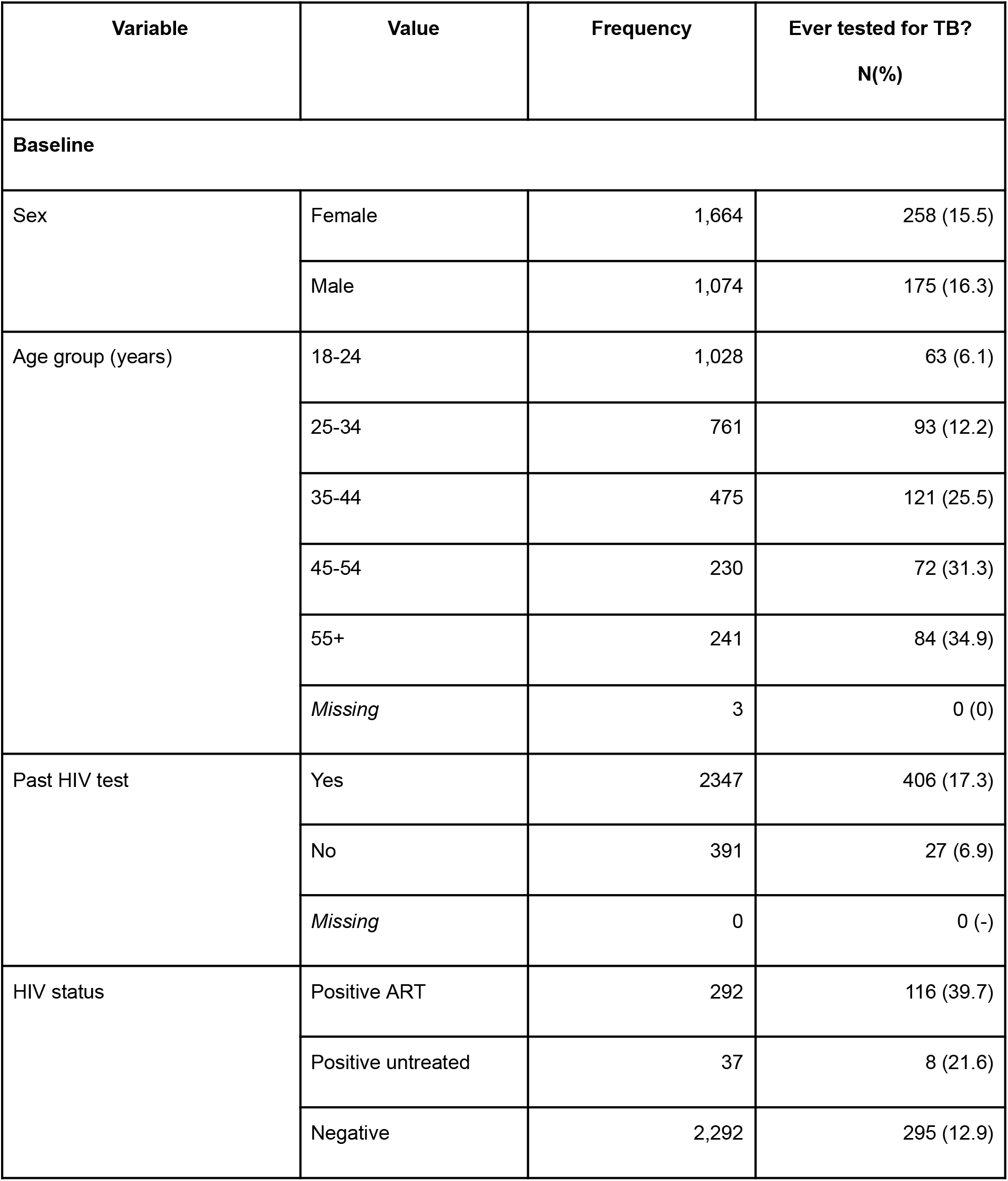
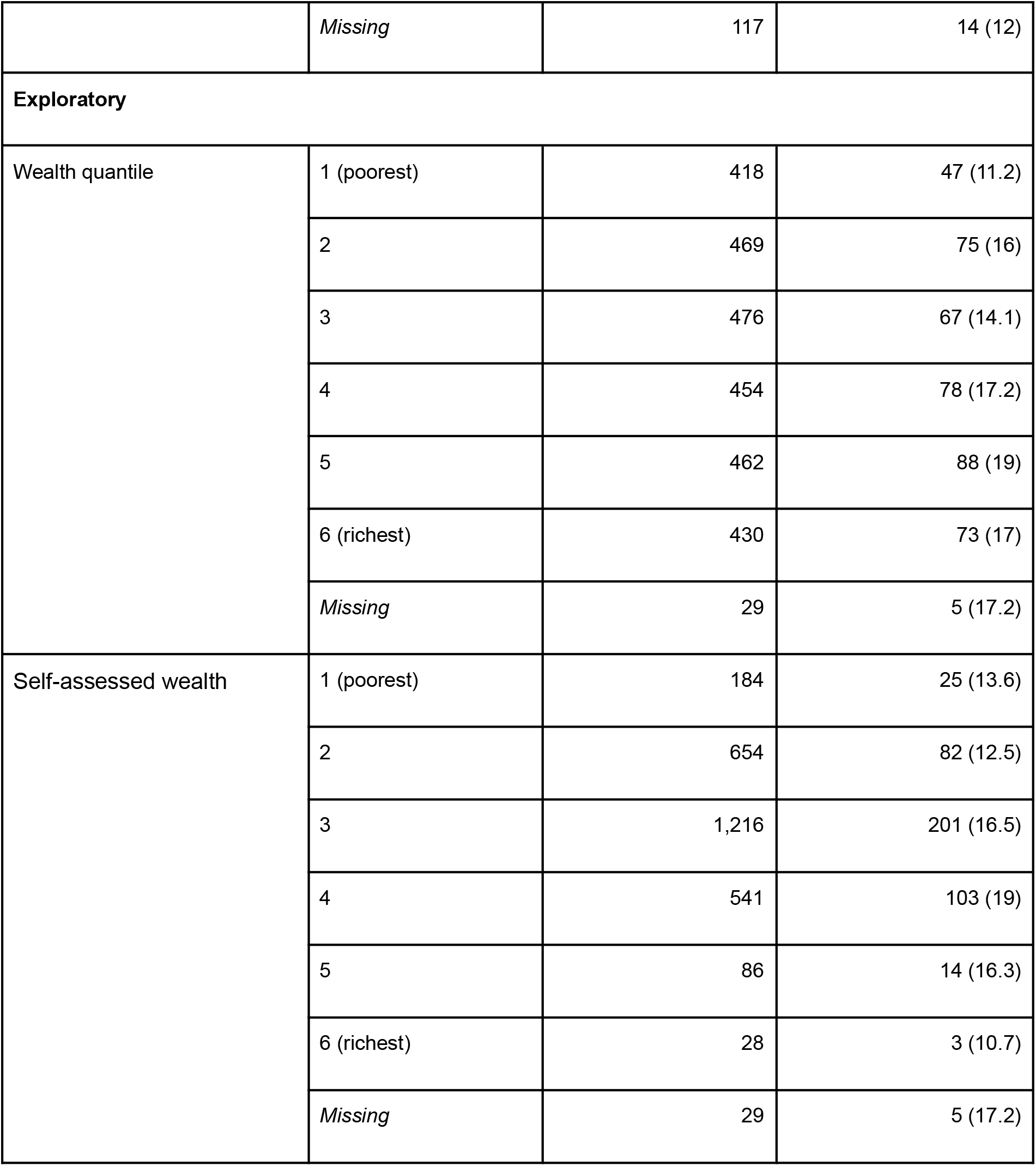
The distribution of key characteristics across the surveyed population, with the percentage reporting previous testing for TB (by sputum sample or chest X-ray).

| Variable | Value | Frequency | Ever tested for TB?<br>N(%) |
| --- | --- | --- | --- |
| **Baseline** |  |  |  |
| Sex | Female | 1,664 | 258 (15.5) |
|  | Male | 1,074 | 175 (16.3) |
| Age group (years) | 18-24 | 1,028 | 63 (6.1) |
|  | 25-34 | 761 | 93 (12.2) |
|  | 35-44 | 475 | 121 (25.5) |
|  | 45-54 | 230 | 72 (31.3) |
|  | 55+ | 241 | 84 (34.9) |
|  | *Missing* | 3 | 0 (0) |
| Past HIV test | Yes | 2347 | 406 (17.3) |
|  | No | 391 | 27 (6.9) |
|  | *Missing* | 0 | 0 (-) |
| HIV status | Positive ART | 292 | 116 (39.7) |
|  | Positive untreated | 37 | 8 (21.6) |
|  | Negative | 2,292 | 295 (12.9) |
|  | *Missing* | 117 | 14 (12) |
| **Exploratory** |  |  |  |
| Wealth quantile | 1 (poorest) | 418 | 47 (11.2) |
|  | 2 | 469 | 75 (16) |
|  | 3 | 476 | 67 (14.1) |
|  | 4 | 454 | 78 (17.2) |
|  | 5 | 462 | 88 (19) |
|  | 6 (richest) | 430 | 73 (17) |
|  | *Missing* | 29 | 5 (17.2) |
| Self-assessed wealth | 1 (poorest) | 184 | 25 (13.6) |
|  | 2 | 654 | 82 (12.5) |
|  | 3 | 1,216 | 201 (16.5) |
|  | 4 | 541 | 103 (19) |
|  | 5 | 86 | 14 (16.3) |
|  | 6 (richest) | 28 | 3 (10.7) |
|  | *Missing* | 29 | 5 (17.2) |

Most respondents self-identified at moderate levels of poverty as opposed to the extremes, with the third step most commonly selected. TB testing was slightly more common among residents of more affluent households compared to less affluent (according to both the measured and self-assessed wealth scales; *Table 1*), and among those who reported knowing someone who has recently tested for or died from TB. Trends across variables appeared broadly similar for the outcomes of ever testing and testing within the past twelve months.

#### Testing history by neighbourhood

An average of 15% of respondents per survey cluster (neighbourhood) reported ever having undergone TB testing (range 0% to 27.5%) (*Figure 2 (A)*). Stratifying by type of TB investigation, greater variability was observed between clusters in the proportion who reported having had a chest X-ray than providing a sputum sample (*Figure 2 (B)*), which is notable given implementation by the National TB Programme and NGO partners of roadside active case-finding delivered by mobile van.

**Figure 1:**
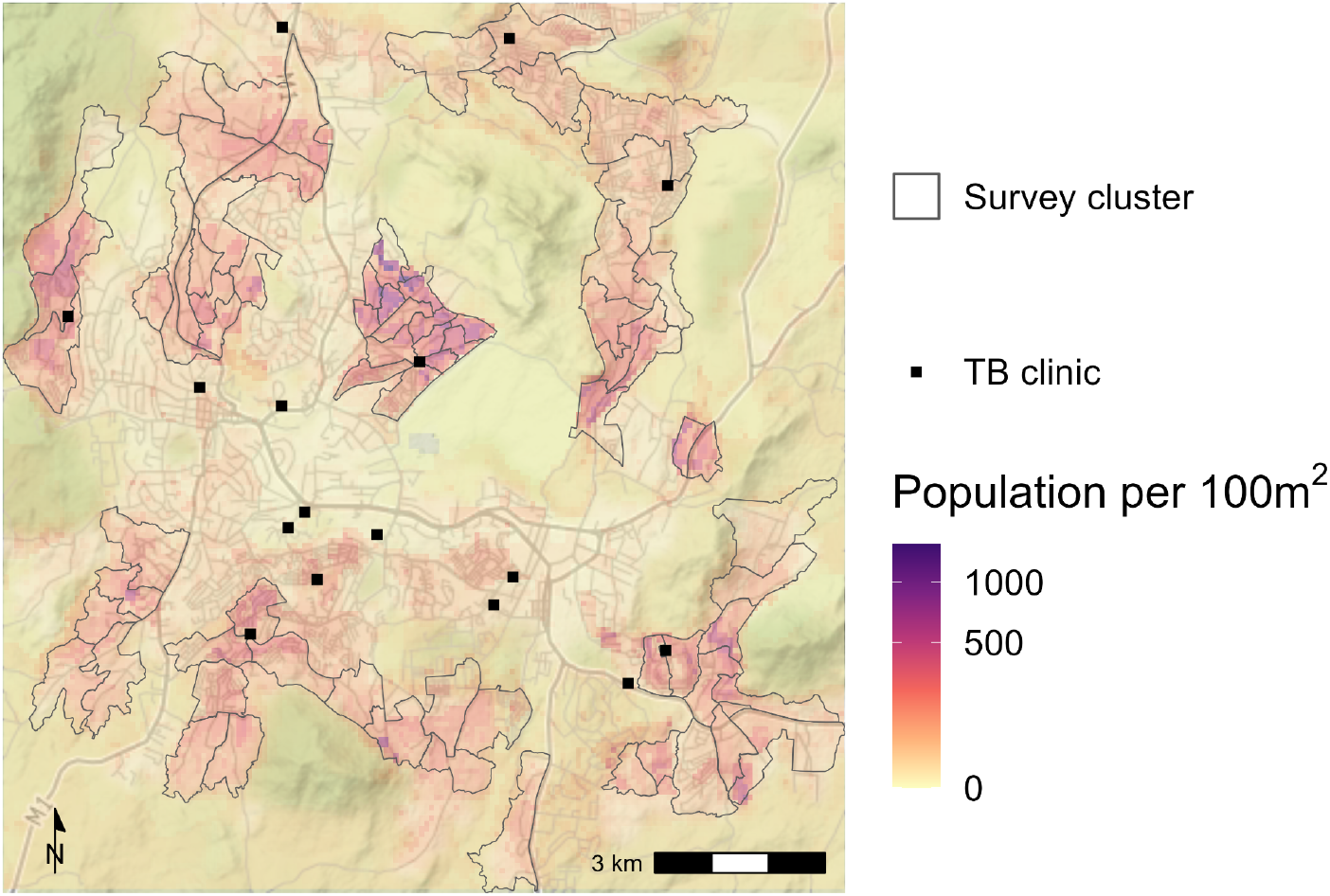
SCALE TB prevalence survey clusters defined across areas of high population density and TB burden in Blantyre City. For illustrative purposes, 100m^2^ pixel population estimates from WorldPop (29) are mapped underneath the cluster boundaries, and locations of TB clinics are indicated in black. For the analysis, however, cluster populations are taken from the MLW census conducted in 2015.

**Figure 2:**
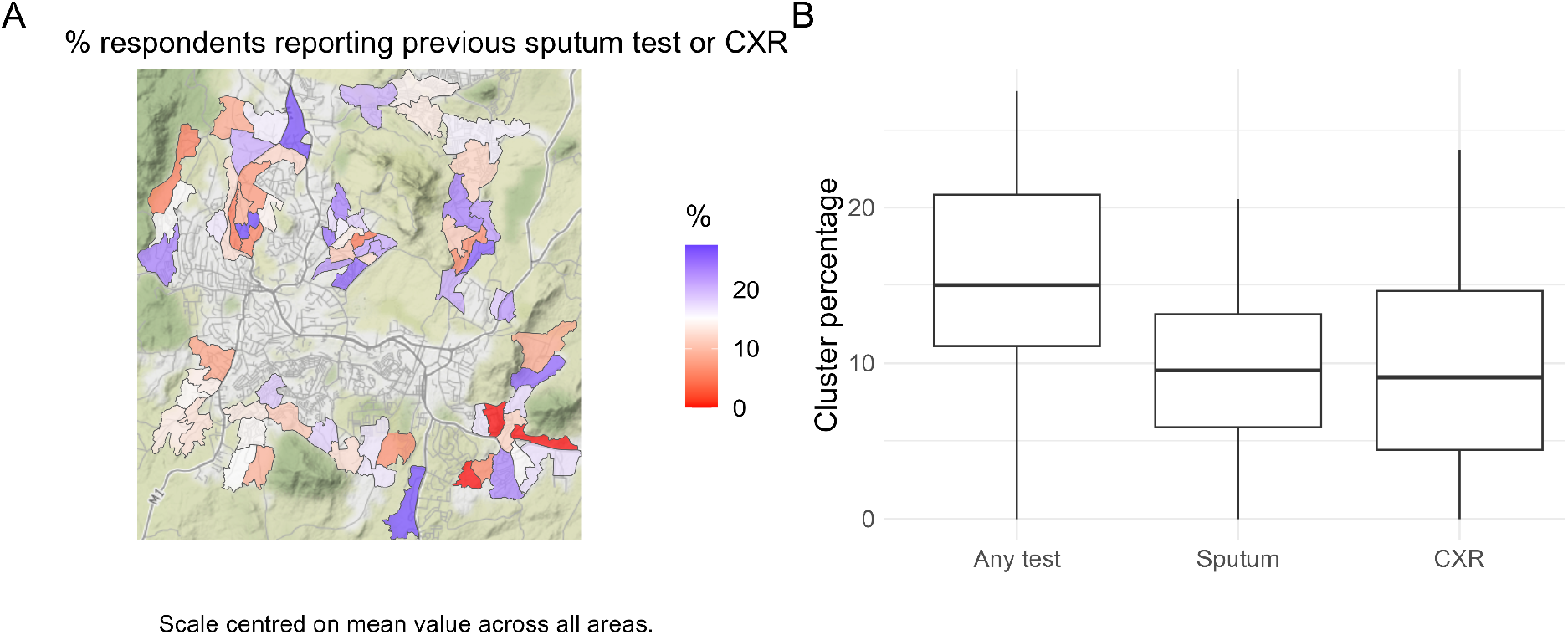
**(A)** Percentage prevalence of self-reported TB testing history (by sputum sample or chest X-ray) across the 72 survey clusters. Locations of TB clinics are indicated in black. **(B)** Distribution of observed cluster percentages by type of test (any, sputum or X-ray). There was greater variation across clusters in the percentage of respondents reporting previous chest X-ray than sputum sample.

#### Poverty by cluster

The distribution of scores from the proxy means test was bimodal, due to the strong influence of the head of household’s education level in the underlying model; the head of household having a higher education qualification substantially reduced the odds of the household living below the poverty line (Figure 3 (A)). However, cluster means of these scores and of self-assessed wealth show broad agreement (Figure 3 (B)).

**Figure 3:**
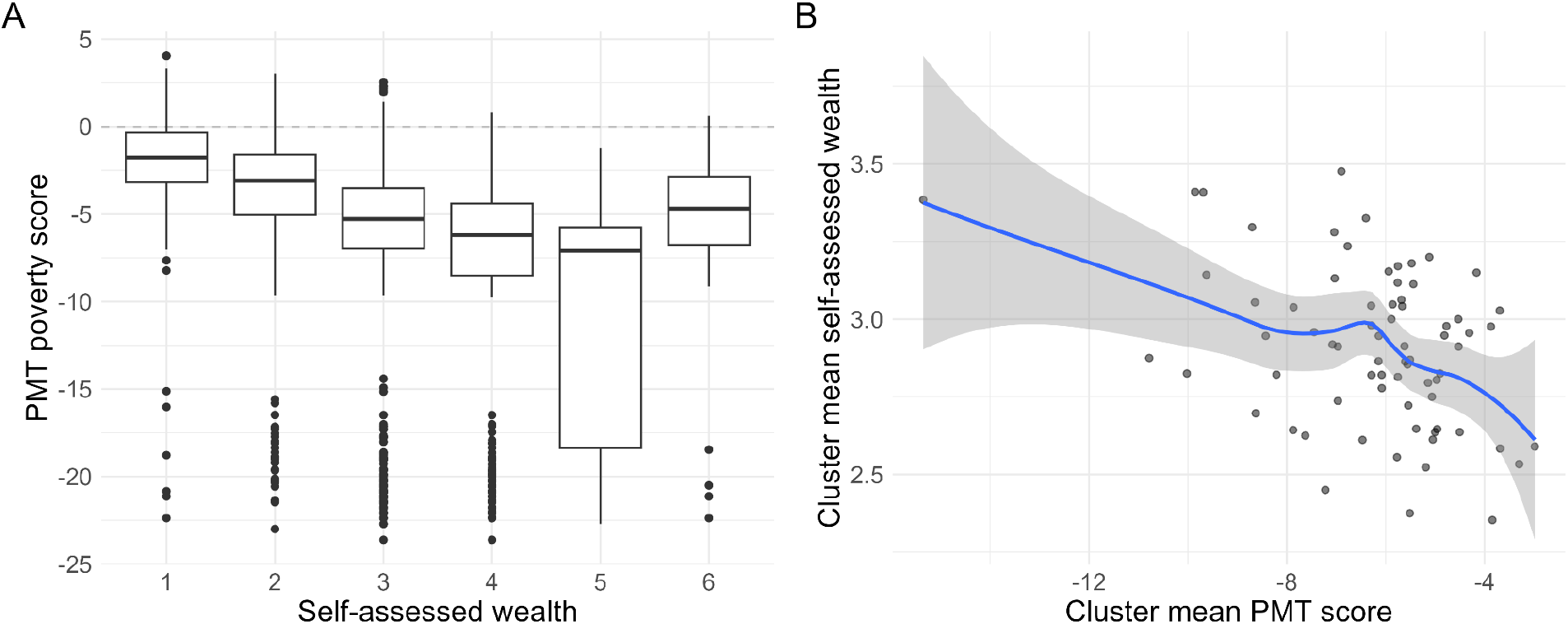
**(A)** The distribution of calculated poverty scores from the proxy means test for each level of self-assessed wealth (1 = poorest, 6 = richest). The bimodality of the overall distribution is reflected in the clusters of “outliers” at much lower values. **(B)** At the survey cluster level, mean self-assessed wealth and PMT poverty scores are broadly in agreement, but exhibit a lot of noise. Note that less negative scores from the PMT reflect greater poverty.

A slightly higher percentage of respondents self-identified at the lowest level of wealth (7%; 95% binomial CI [6, 8]) than were predicted to be below the poverty line according to their proxy means test responses (5%; 95% CI [4, 6]). Visually, self-assessed wealth appeared to reflect smoother trends between clusters than PMT scores, with greater similarity between neighbouring clusters (*Figure 4*).

**Figure 4:**
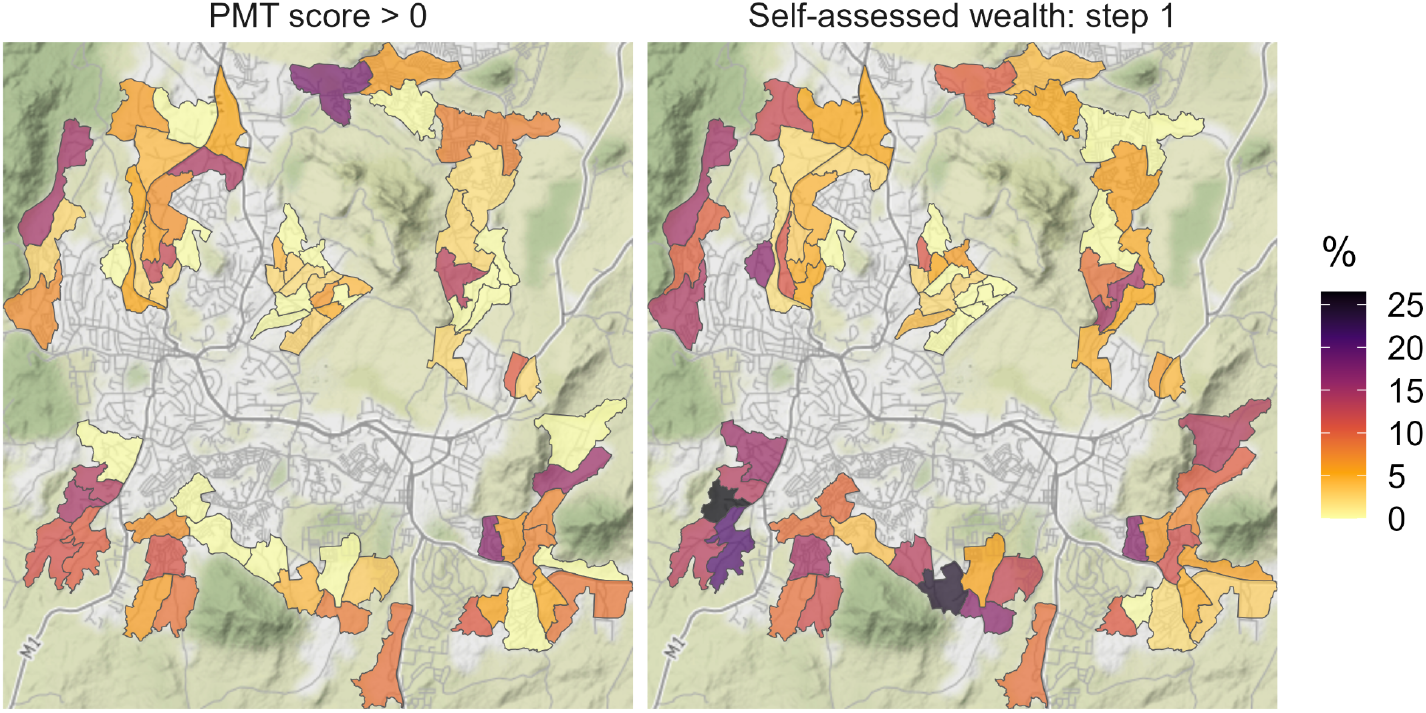
Summary of the two measures of poverty by survey cluster. On the logit scale, a PMT score greater than zero indicates that more likely than not the household lives below the poverty line. The proportion of households per cluster for which PMT > 0 (left) is compared to the proportion who self-identified on the lowest level of wealth (right).

### Baseline model fit

Graphical assessment suggested differing age trends between men and women with respect to reported TB testing history (*Supplementary Figure S4*), therefore an interaction term was included in the baseline model specification. This yielded a small decrease in LOOIC from a model without interaction (2043 (standard deviation/SD 61.8) vs 2048 (61.6), respectively).

The baseline fit clearly demonstrated that older age was associated with greater odds of reporting having ever undergone testing for TB, with a larger magnitude of effect for men compared to women (OR for age ≥ 55 vs < 25 years = 4.0 [2.33, 6.67] for women; 9.3 [5.50, 15.41] for men) (*Figure 5 (A)*). The impact of routine testing among known HIV-positive people was also evident (OR of ever testing for HIV-positive people = 2.72 [2.04, 3.60] compared to HIV negative). Unknown HIV status was considered as a missing value and hence observations excluded from complete cases analysis; however, as a sensitivity analysis it was found that including it as a factor level to be estimated had negligible effect on the coefficient for HIV-positive (OR 2.7 [2.03-3.55]).

**Figure 5:**
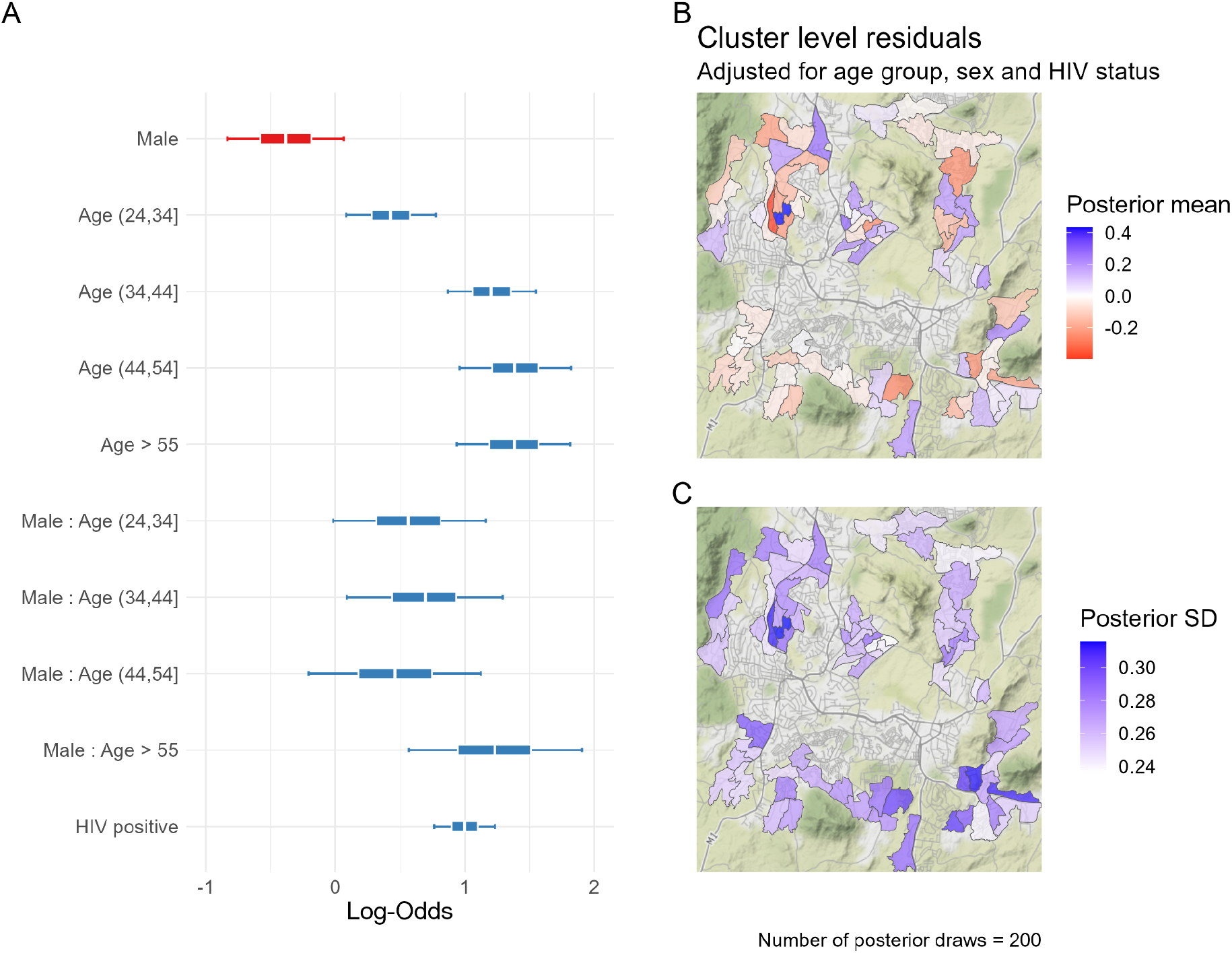
**(A)** Fixed effect estimates from baseline model fit; a log-odds ratio greater than 1 indicates a higher chance of having been tested relative to the baseline for that variable **(B)** Variation across clusters in the cluster-level random effect (posterior mean); blue and red indicate where testing prevalence in the cluster is generally higher or lower than expected, respectively, having adjusted for each respondent’s age, sex and HIV status. **(C)** Posterior standard deviation (SD) of the cluster-level random effect.

The cluster-level random effects demonstrated that heterogeneity remained between clusters after accounting for variation in the distributions of individual characteristics (posterior SD 0.3 [0.08 - 0.51]) (*Figure 5 (B)*). Some individual clusters stood out in contrast to their neighbours in the centre-north and north-east of the city, and overall there appeared to be no strong pattern across clusters.

### Explaining residual cluster-level variation

For the proxy means test, lower odds of ever having been tested for TB was associated with greater log-odds of the individual’s household living below the poverty line. The negated continuous score was associated with a borderline significant positive trend, and those in the quantile least likely in poverty were estimated to have significantly higher odds of TB testing than those in the poorest quantile (*Table 2; Supplementary Figure S6 (A)*). Odds were also significantly higher among those who *self-identified* at the 3rd and 4th highest steps of wealth - relative again to the poorest - as was observed in unadjusted comparison.

**Table 2:** Comparison of fixed effect estimates from three models incorporating poverty through three different metrics: PMT poverty score (predicted probability of living on < $2/day), quantiles of poverty score, and self-assessed wealth on a six-point scale. Including PMT poverty score yielded the greater reduction in the leave-one-out information criterion (LOOIC).

| Variable | Estimate | Change in LOOIC from baseline |
| --- | --- | --- |
| PMT score quantile: 1 (poorest) | Reference | -5.4 |
| 2 | 1.4 (1.01-2.00) |  |
| 3 | 1.2 (0.86-1.76) |  |
| 4 | 1.9 (1.39-2.69) |  |
| 5 | 1.8 (1.33-2.60) |  |
| 6 (richest) | 1.7 (1.21-2.83) |  |
| Self-assessed wealth: 1 (poorest) | Reference | -1.3 |
| 2 | 1.1 (0.74-1.73) |  |
| 3 | 1.6 (1.11-2.47) |  |
| 4 | 1.8 (1.21-2.83) |  |
| 5 | 1.5 (0.81-2.78) |  |
| 6 (richest) | 1.1 (0.34-2.99) |  |

Despite the significance of poverty measure coefficients, neither improved the overall fit of the model to a substantial degree (*Table 2*). Moreover, neither measure appeared to explain a greater extent of the cluster-level variation than the baseline model, having negligible impact on the estimated SD of the random intercept (*Supplementary Figure S6 (B*)).

Multiple imputation of missing values in HIV status and the two household poverty variables showed negligible impact on estimated coefficients compared to the presented complete case analysis (*Supplementary figures S7 and S8*).

### Comparison with prevalence-to-notification ratios

The cluster-level residuals from either the baseline or poverty-adjusted models showed no visible association with previously estimated ratios between prevalence and case notifications (*Supplementary figure S9*), although these were limited by low numbers of prevalent TB cases, as detailed above.

## Discussion

The main findings from this analysis of questionnaire data from a pre-intervention survey for undiagnosed infectious TB in Blantyre, Malawi, was that 16.0% (95% CI [14.6%, 17.4%]) of adults reported either having submitted sputum or having had a chest X-ray at some time in the past - likely to have represented past TB testing. By comparison, 85.7% of survey participants reported at least one previous HIV test. Past TB testing increased with age, but even among the oldest participants (55 years and older) only 34.9% had ever tested for TB. Most TB testing was recent, likely reflecting recent scale-up of TB case-finding efforts by the National TB and HIV Programmes over the last decade, including decentralisation of TB testing and treatment, more intensive implementation of systematic screening nationally in outpatient attendees, people living with HIV, and prisoners (21).

In urban areas including Blantyre, case-finding efforts have included community-wide active case-finding by mobile digital X-ray vans and door-to-door symptom screening following results of a 2013-14 National TB Prevalence survey showing ∼1% of urban adults with undiagnosed infectious TB (5,22). Reported TB testing rates varied substantially between neighbourhood clusters, even after adjusting for local age-sex distributions and HIV-positive status.

Prior TB testing was associated with household-level poverty, whether measured by self-assessment or by a proxy means test, yet neither measure explained the residual variation by cluster. This suggests that poverty was not a predominant driver of spatial variation in TB testing in this population, and could reflect the complexity and heterogeneity of barriers to accessing and engaging with testing *within* clusters as opposed to between clusters (4). The low coverage of life-time testing suggests a need to focus more efforts on self-reported TB testing data, as has been successfully used with HIV regionally, in order to identify and target underserved subgroups and communities as we work towards EndTB targets.

Estimated prevalence of adult pulmonary TB was 452 (312-593) per 100,000 adults nationally and 1,014 (486 to 1,542) for urban adults (> 15 years) in the 2013-14 Malawi national prevalence survey (5,22), but had fallen substantially by the 2019-20 SCALE pre-intervention survey to 180 per 100,000 adults in Blantyre (8). The 2013-14 survey qualified Malawi as a high disease burden country, with the urban areas meeting the threshold for which general population active case-finding is conditionally recommended by WHO, as implemented by the National Programme and implementing partners. A systematic review by Burke *et al* (23) discussed the potential importance and impact of active case-finding if delivered with sufficient intensity. Our analysis, however, suggests that reach and coverage may have been inconsistently achieved between otherwise comparable communities across Blantyre.

Implementation in Malawi has used symptom-based screening algorithms, so that only participants reporting TB symptoms progress to chest X-ray and/or sputum examination. This approach reduces numbers needed to test, and but has considerably lower sensitivity than alternative systematic screening approaches, such chest X-ray or sputum testing of all individuals irrespective of symptoms (24), and is likely to be contributing to the relatively high proportion of adult residents in Blantyre who have never been investigated with chest X-ray or sputum. However, this alone is unlikely to explain all of the heterogeneity seen in this study, especially regarding the lower coverage of testing for the most poor in any given community. Increasing equity of access to care should be a goal of national TB programmes, and with monitoring of the reach and coverage of TB testing a potentially important source of data to ensure certain groups are not under-represented. As with case-notification data, national programmes could map coverage of TB testing events and identify under-testing prospectively if geospatial data are available for each tested patient, for example as demonstrated in (10).

We did not find that the observed variation in past TB testing was readily explained by neighbourhood-level poverty; rather, the most poor individuals in all neighbourhoods were the least likely to have used Malawi’s free health services for TB diagnostic investigation. A more in-depth analysis is beyond the scope of this manuscript but could investigate how motivations and behaviours may vary for those in extreme poverty as well as across the city. We also did not, for example, assess travel distance to TB clinics as a potential barrier. In previous work, Khundi et al. found distance to be a risk factor for TB case fatality among patients notified at the central hospital in Blantyre (25).

After accounting for individual HIV status and poverty, variation in past TB testing may well act at the clinic or practitioner level rather than across survey clusters. However, residual cluster-level variation did not show a strong trend of correlation between neighbouring clusters.

Survey cluster boundaries were based on community health worker catchment areas, which in turn are derived from health clinic catchment areas. As such, we would have expected spatial correlation within the same catchment area if an important source of variation was at the clinic level.

Our socioeconomic analysis used two measures of poverty: a proxy means test based on a 15-point questionnaire derived from a recent extensive World Bank Integrated Household

Survey which defined poverty based on over 200 questions on income, assets, production and consumption, and a single self-rated question also drawn from the Integrated Household Survey. In this study the two measures were broadly but not fully consistent. Self-assessment of wealth status is quick and easier to collect than deriving and then asking the multiple components of the proxy means test, but has no frame of reference and hence may not be comparable between settings. Self-rated wealth in our survey also resulted in imbalanced categories, with more individuals avoiding the extremes of the scale. Within a constrained population (as sampled here), our data and the relationship between poverty and past health seeking behaviour adds support to self-assessment as a useful relative measure of poverty that may, for example, capture more subtleties than a quantitative score based on a limited number of fixed indicators. It also does not need updating over time as relevant assets and indicators change.

A systematic review found that explicitly spatial analyses of TB epidemiology have rarely accounted for undiagnosed cases (26). Rood et al. (27) used rates of TB testing as a covariate when modelling TB incidence at a local geographic level. Shaweno et al. (28) demonstrated a method to account for underdiagnosis using routinely collected data in Ethiopia and conclude a substantial number of missed cases, yet did not allow for different rates of diagnosis in different locations or populations. We propose that incorporating data on recent history of testing into analysis of case notifications could be used to make this type of adjustment for under-diagnosis, allowing routine programmes to start to identify regional and national diagnostic barriers.

## Limitations

These data were drawn from a prevalence survey that had a suboptimal participation rate, notably for working age men. This could have biased estimates from the current analysis, for instance, underestimating the role of poverty if the poorest men were also the most likely to have been non-participants and the least likely to have tested for TB in the past. Due to the structure of the model, re-weighting of observations through post-stratification to adjust for this type of bias would require cluster-level estimates of HIV prevalence by both age and sex, which were not available at time of writing. Our definition of past TB testing will have included some misclassification, as individuals may have had a sputum collection or chest X-ray for reasons other than TB. In practice, however, these examinations are rarely conducted for other reasons in Malawi. We also could not explore recency of testing with these data due to low numbers of participants able to specify exactly when their last test was.

## Conclusions

Despite decades of emphasis on TB case-finding and recent revision of global guidance to provide broader support for community-based active case-finding and chest X-ray based screening, few studies have investigated population patterns of prior TB testing as a potentially important marker of programmatic reach and equity. To our knowledge, this is the first analysis of TB testing history measured from a city-wide survey. Although Blantyre has experienced a generalised HIV epidemic and extremely high TB burden, only a small proportion of community residents reported having previously been tested for TB. In spatial regression analysis, neighbourhood-level variation in TB testing was not explained by demographics, HIV positivity or household level poverty, suggesting that access to, and quality of, healthcare provision remains a persistent barrier to TB screening and diagnosis services. This motivates further work to understand how behavioural drivers including opportunity (both social and physical), individual motivation and capacity may contribute to this variability. As TB epidemics in Africa decline and concentrate in vulnerable groups, improving access, acceptability and equity in provision of high quality TB services will be essential to accelerate progress towards TB elimination goals and a greater focus on collection and analysis of TB testing data will be needed.

## Data Availability

All data produced in the present study are available upon reasonable request to the authors.

https://github.com/esnightingale/tb-testing-history

## Acknowledgments

The authors would like to thank Lingston Chiume at the Malawi Liverpool Wellcome Trust (MLW) Clinical Research Programme for their support.

First author Emily S. Nightingale is a research fellow in statistical modelling at the London School of Hygiene and Tropical Medicine. Her research primarily focuses on the spatial epidemiology of infectious diseases, with work on leishmaniasis, TB and COVID-19.

## Ethical statement

Ethical approval for the Sustainable Community-wide Active case finding for Lung hEalth (SCALE) trial (registered trial number: ISRCTN11400592) was granted by the LSHTM Research Ethics Committee (Ref: 16228) and by the University of Malawi College of Medicine Research and Ethics Committee (Ref: P.12/18/2556).

## Funding

The SCALE trial was funded by the Wellcome Trust (200901/Z/16/Z).

## Supplementary Materials

## Notes

### Competing Interest Statement

The authors have declared no competing interest.

### Author Declarations

Ethics committee of the London School of Hygiene and Tropical Medicine gave ethical approval (Ref: 16228) for this work as part of the Sustainable Community-wide Active case finding for Lung hEalth (SCALE) trial (registered trial number: ISRCTN11400592). Ethics committee of the University of Malawi College of Medicine gave ethical approval for this work as part of the Sustainable Community-wide Active case finding for Lung hEalth (SCALE) trial (Ref: P.12/18/2556).

