## Supplementary Materials for "Community-level variation in TB testing history: analysis of a prevalence survey in Blantyre, Malawi"

#### A Target population and sample representativeness

Total sex-stratified populations within the survey clusters were estimated from a study census conducted in 2015 by the Malawi-Liverpool-Wellcome Programme (MLW) (7), and the Malawi national censuses conducted in 2008 and 2018. Methods used to estimate cluster populations at the time of the prevalence survey have been described previously (5). We summarised characteristics of study participants, and compared the sex and age distribution within the sample of extended questionnaire respondents to that of the total population of the study clusters.

The sample of extended survey respondents consisted of a greater proportion of women than the total resident population of the survey clusters (60.8% vs. 29.4% female, respectively) and also than the estimated resident population of sampled households (52%) (*Supplementary Figure S1 (A)*). This difference appeared to primarily arise from underrepresentation of working-age men (*Supplementary Figure S1 (B)*), who may have been reported as residents by another household member but not present to participate in the individual survey.

#### B Definition of HIV status

All adult residents participating in the survey were asked about their HIV status from any previous testing, and also offered Oraquick (Orasure) and Determine (Alere) rapid testing in parallel during attendance of TB screening. Overall, 75% (2044/2738) of those participating in the extended survey completed HIV testing. Screening results were preferred and self-reports only used if this was not available. For those attending the testing site, self-reported positive status from a previous test and positive rapid test results were confirmed with Uni-Gold (Trinity Biotech).

Patients who self-reported as positive from a previous test result, self-reported ART use or attendance of an ART clinic, or who tested positive upon screening were classified as being HIV-positive. Additionally, participants who reported ART use or attendance of an ART clinic were classified as being “HIV-positive on ART”. Patients testing HIV-negative during screening or reporting a previous negative HIV result, and not using ART, were classified as “HIV negative”. Patients who had unknown or missing responses and unknown or inconclusive screening results were classified as having “unknown” HIV status.

#### C Model specification

Weakly informative priors were set on all parameters. A normal prior with mean -2 and standard deviation (SD) 2 was specified for the global intercept, assuming that overall a minority of participants would have had prior testing for TB. Fixed effect coefficients were assigned student’s t priors with mean 0, 3 degrees of freedom and scale 5. The standard deviation of the cluster level random intercepts was assigned a half-Cauchy prior with mean 0 and scale 1. Prior predictive plots indicated that this structure replicated an appropriate distribution of outcomes (*Supplementary figure S5*).

For individual $i$ resident in cluster $j$, with baseline covariate values $X_i$, the general model structure is therefore defined as follows:

$$Y_{ij} \sim Bernoulli(p_{ij})$$

$$logit(p_{ij}) = u_j + \beta X_i$$

$$u_j \sim Normal(\beta_0, \sigma)$$

$$\beta_0 \sim Normal(-2, 2)$$

$$\beta_k \sim T(0, 3, 5), \quad \beta_k \in \{\beta_1, ..., \beta_K\}$$

$$\sigma \sim Half - Cauchy(0, 1)$$

The model was fitted using Hamiltonian Monte Carlo with No U-Turn Sampling (NUTS) via the R package *brms* as an interface to Stan (24,25). The joint posterior distribution was estimated through four chains each of 2,000 iterations (1,000 burn-in). Posterior estimates for the coefficients of the fixed covariate effects and the standard deviation of the random intercepts are presented with 95% Credible Intervals (CrI).

### Supplementary Figures

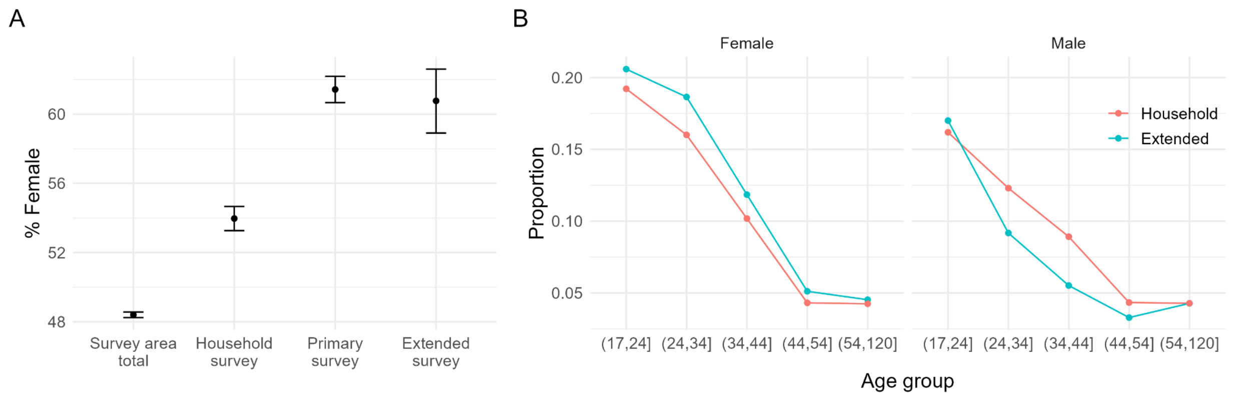


***Figure S1:*** ***(A)*** *Proportion of female residents (with exact 95% confidence intervals) according to the total cluster population, the household survey, primary individual survey and extended survey.* ***(B)*** *Age distribution of males and females within the sample populations reported through the household survey, and within the extended individual survey.*

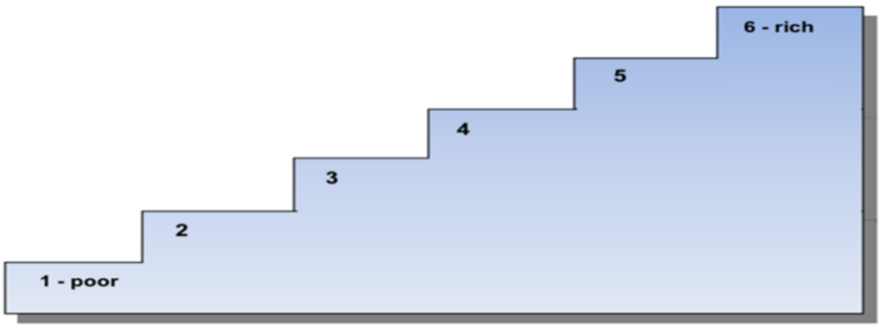


***Figure S2:*** *Illustration used in the household questionnaire for self-assessment of wealth on six steps.*

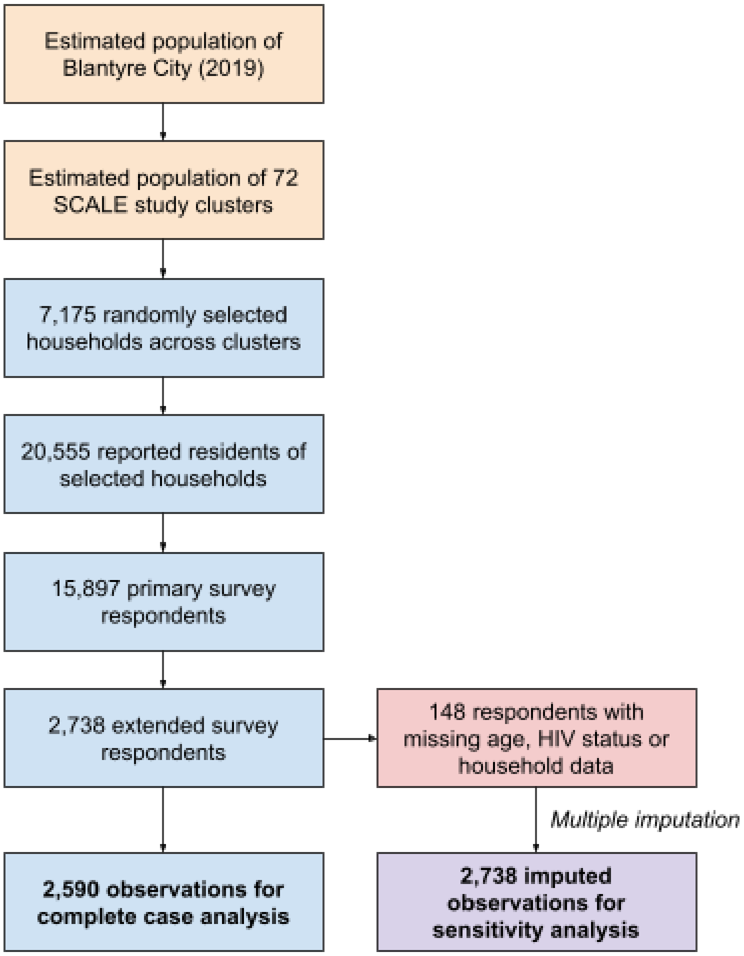


***Figure S3:*** *Flowchart of sampling process and definition of the primary analysis dataset.*

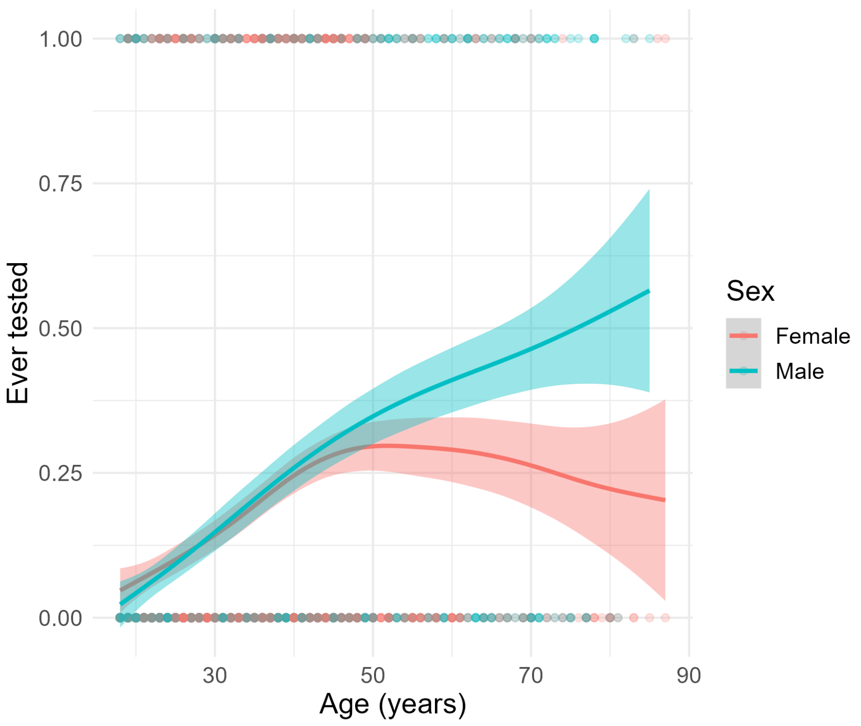


***Figure S4:*** *Graphical assessment of interaction between age and sex, with respect to reported ever testing. Specifically, the proportion increases approximately linearly with age among men, while the trend among women diverges from this at around 40 years.*

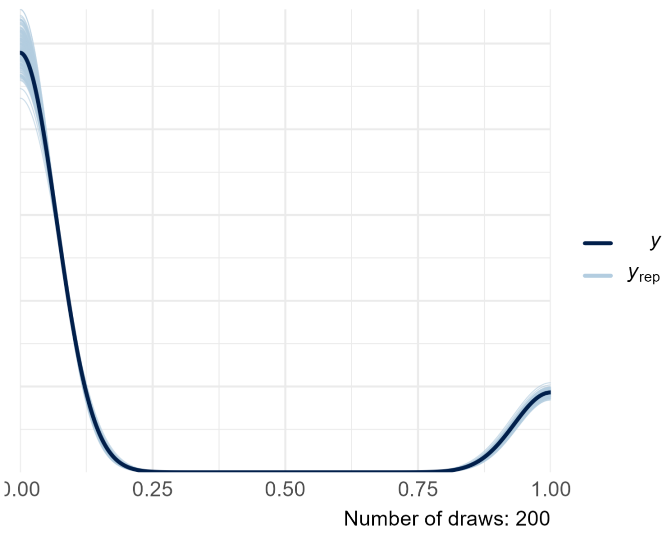


***Figure S5:*** *Prior predictive distribution from the null, intercept-only model (200 draws).*

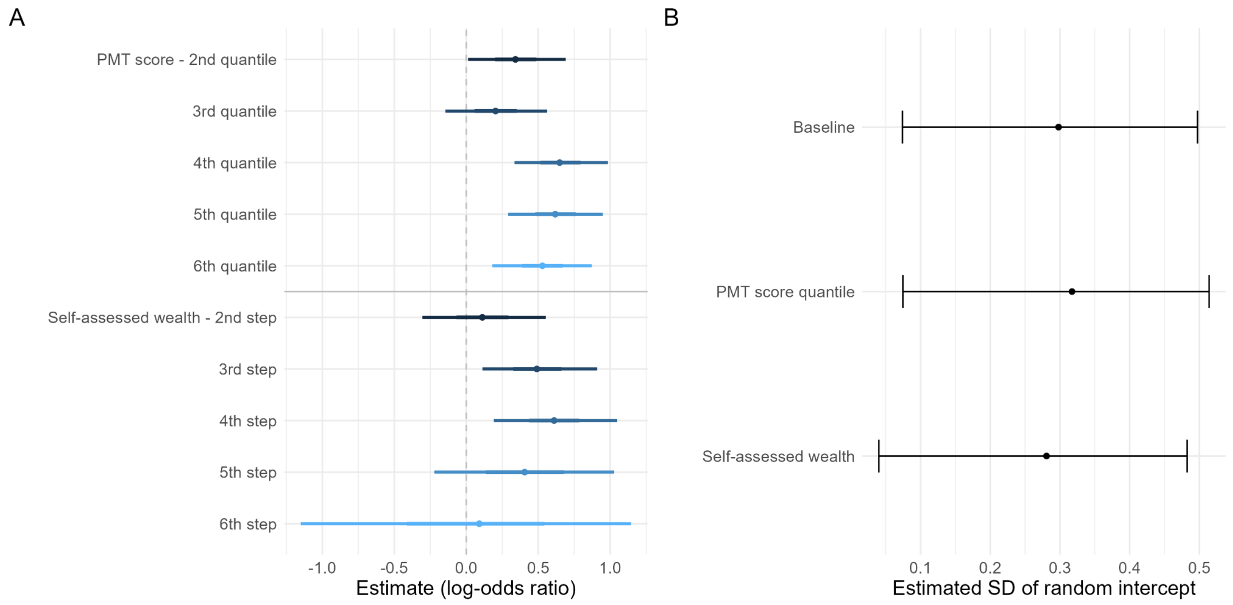


***Figure S6: (A)*** *Posterior estimates of fixed effect coefficients for the two poverty measures. Each was added to the baseline model independently and then compared by LOOIC.* ***(B)*** *Posterior estimates for the SD of the cluster-level random intercepts.*

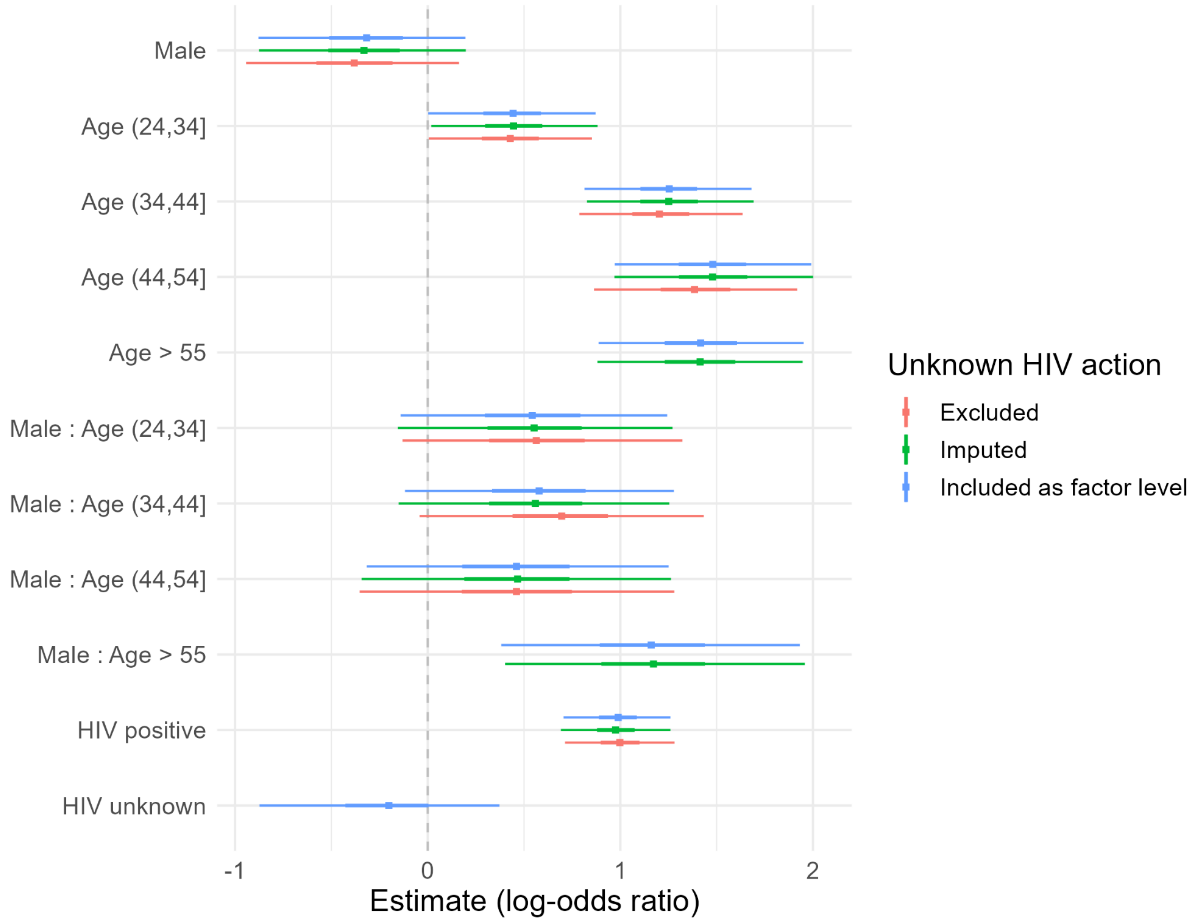


***Figure S7:*** *Comparison of fixed effect estimates with different actions to address unknown HIV status: exclusion, multiple imputation and estimation as an additional factor level. 50% and 95% credible intervals are shown around the point estimate.*

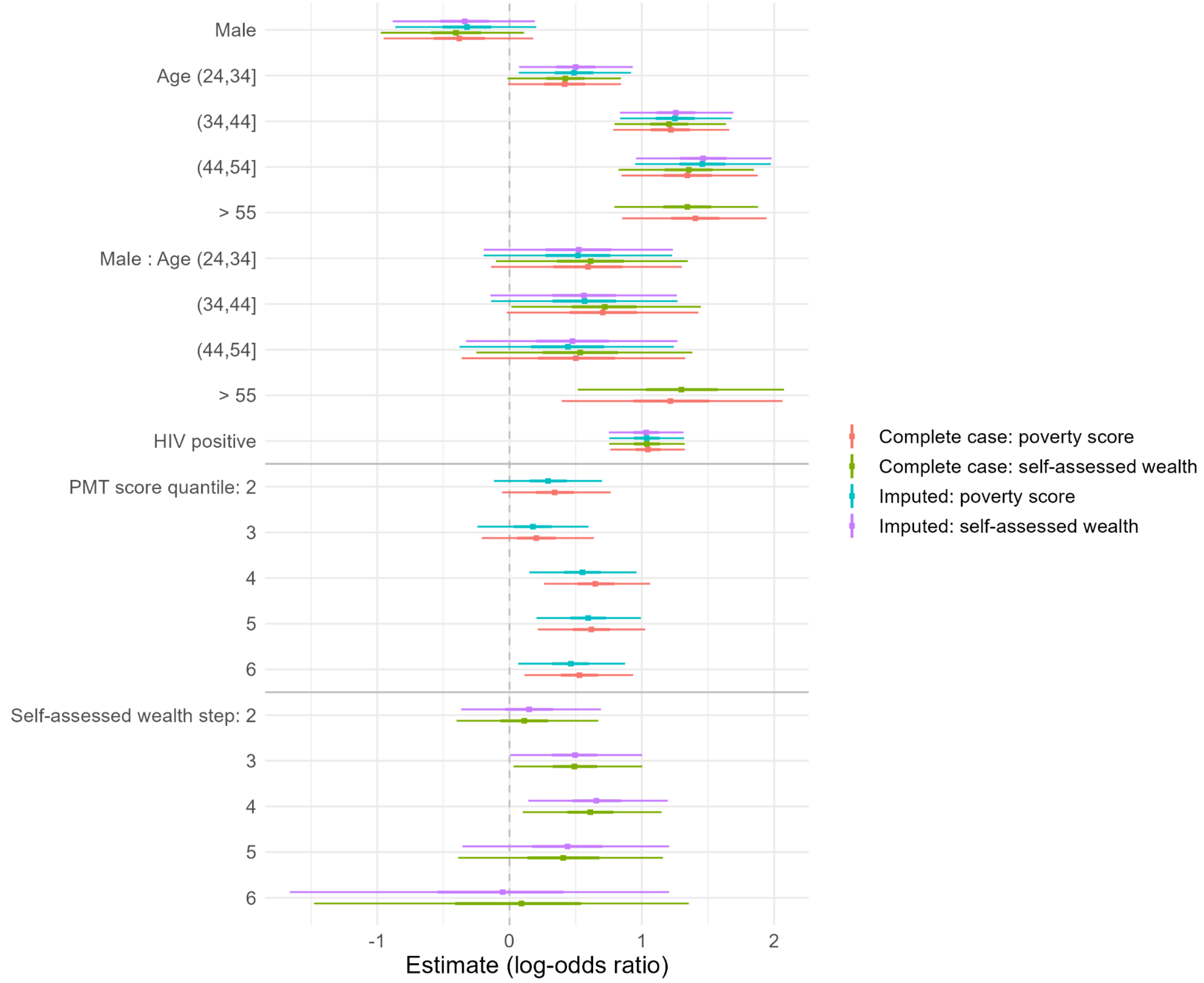


***Figure S8:*** *Comparison of fixed effect estimates from complete case analysis with multiple imputation of unknown HIV status and household poverty variables.*

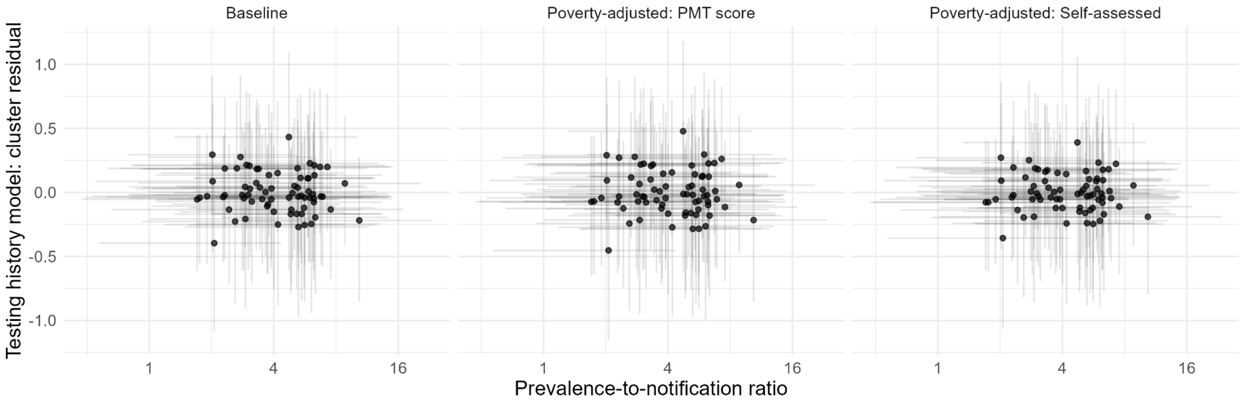


***Figure S9:*** *Fitted cluster-level residuals (posterior mean and 95% CrI) from the baseline model plotted against the prevalence-to-notification ratios (PNRs) estimated as part of a previous analysis of the SCALE data (6). These PNRs reflect the disparity between TB prevalence as estimated from the SCALE survey and observed case notifications, hence the extent of underdiagnosis per cluster. There is no apparent relationship between unexplained cluster-level variation in past TB testing and the estimated extent of underdiagnosis.*
